# Prevalence of Meningitis and Pneumonia among Neonates Treated at Public Hospitals in Ethiopia: A Systematic Review and Meta-Analysis

**DOI:** 10.64898/2026.07.16.26358228

**Authors:** Hermella Negash Woldearegay, Maedot Sebsibe Tebeka, Saeed Omer Saeed Osman

## Abstract

**Background:** Neonatal meningitis and pneumonia are important causes of morbidity and mortality among hospitalized newborns in low-resource settings. Numerous single-centre studies have been conducted in Ethiopian public hospitals, yet no quantitative synthesis of neonatal meningitis prevalence previously existed, and the depth of the pneumonia-specific evidence base was undocumented. This review aimed to estimate the pooled prevalence of neonatal meningitis, and to establish the state of the evidence base for neonatal pneumonia, among neonates treated at public hospitals in Ethiopia.

**Methods:** A systematic search of PubMed/MEDLINE, PubMed Central, PLOS, BioMed Central, Frontiers, Taylor & Francis Online, and institutional repositories was conducted without date restriction. Cross-sectional and retrospective studies reporting a standalone prevalence of neonatal meningitis or neonatal pneumonia among neonates treated at Ethiopian public hospitals were eligible. Methodological quality was appraised using the JBI Critical Appraisal Checklist for prevalence studies. A random-effects meta-analysis pooled meningitis prevalence; heterogeneity was quantified with I² and Cochran’s Q.

**Results:** Of 26 full-text articles assessed for eligibility, four studies (N = 3,522 neonates) met inclusion criteria for neonatal meningitis; none met inclusion criteria for a standalone neonatal pneumonia prevalence outcome. The pooled prevalence of neonatal meningitis was 6.26% (95% CI: 2.81–13.35%; I² = 96.6%), with individual study estimates ranging from 1.73% to 19.30%. A sensitivity analysis restricted to the three studies using a “suspected-meningitis” denominator yielded a pooled prevalence of 4.23% (95% CI: 1.92–9.05%). No eligible primary study reported neonatal pneumonia prevalence as a standalone, separately ascertained outcome across the accessible literature, pneumonia is consistently subsumed within composite “neonatal sepsis” case definitions.

**Conclusion:** An estimated 1 in 16 to 1 in 24 neonates tested for suspected meningitis at Ethiopian public hospitals had a culture-confirmed or clinically diagnosed case, with wide variation across settings and case-ascertainment methods. A previously undocumented evidence gap exists for standalone neonatal pneumonia prevalence in Ethiopia. Future primary studies should report pneumonia as a distinct, separately ascertained neonatal outcome to enable future quantitative synthesis.

## 1. Introduction

Globally, an estimated 2.4 million neonates die each year, with the majority of these deaths concentrated in sub-Saharan Africa and South Asia. Infections principally sepsis, meningitis, and pneumonia together with complications of prematurity and intrapartum-related events (birth asphyxia), account for the great majority of these deaths. In Ethiopia, despite measurable declines in the neonatal mortality rate over the past two decades, neonatal deaths now represent a growing proportion of all under-five deaths as post-neonatal child mortality has fallen faster; infection-related causes, including sepsis, meningitis, and pneumonia, remain among the leading contributors alongside prematurity and perinatal asphyxia.

Neonatal meningitis, an inflammation of the meninges most often caused by bacterial pathogens acquired vertically from the maternal genital tract or horizontally within the healthcare environment, carries a disproportionately high burden of mortality and long-term neurodevelopmental sequelae (hearing loss, seizure disorders, cognitive impairment) when compared with meningitis at older ages. Ethiopia lies within, or adjacent to, the African “meningitis belt”, a region of historically elevated bacterial meningitis burden. Single-centre Ethiopian studies have reported the prevalence of neonatal meningitis among neonates undergoing diagnostic work-up ranging from below 2% to nearly 20%, a range wide enough to suggest that pooling with explicit attention to sources of heterogeneity would meaningfully advance the evidence base.

Neonatal pneumonia is similarly recognized as a major contributor to neonatal respiratory morbidity and mortality, arising from congenital or transplacental or intrapartum-acquired or postnatal nosocomial or community-acquired) infection. In global and regional estimates, pneumonia is consistently named among the leading infectious causes of neonatal death. However, unlike meningitis for which Ethiopian hospitals routinely perform and report a discrete diagnostic test such as erebrospinal fluid culture the operational case definition of “neonatal sepsis” used in the overwhelming majority of Ethiopian hospital-based studies explicitly incorporates pneumonia as an unspecified subtype, alongside septicaemia, meningitis, and other invasive infections, without disaggregated case counts. This creates a structural barrier to synthesizing pneumonia-specific prevalence, distinct from any true absence of the underlying clinical problem.

To our knowledge, no prior systematic review has

- Produced a pooled, quality-appraised estimate of neonatal meningitis prevalence specific to Ethiopian public hospitals,
- Formally documented the extent of comparable primary evidence for neonatal pneumonia in this setting. Addressing both conditions within a single review allows the second finding to be established with the same systematic rigor as the first, rather than being asserted informally.

### 1.1 Objectives

#### General objective

To determine the pooled prevalence of meningitis and pneumonia among neonates treated at public hospitals in Ethiopia.

#### Specific objectives

● To estimate the pooled prevalence of neonatal meningitis among neonates treated at Ethiopian public hospitals, using a random-effects meta-analysis.
● To determine, through a systematic and transparent search and screening process, whether sufficient primary evidence exists to estimate a pooled prevalence of neonatal pneumonia in the same setting.
● To explore heterogeneity in neonatal meningitis prevalence estimates through sensitivity analysis.
● To appraise the methodological quality of included studies and assess risk of publication bias.

## 2. Methods

This review was conducted with reference to the Preferred Reporting Items for Systematic Reviews and Meta-Analyses (PRISMA) 2020 statement.

### 2.1 Eligibility Criteria

Studies were eligible if they:

i. Enrolled neonates defined as 0–28 completed days of life, or a study-defined equivalent such as <90 days for late-onset meningitis case ascertainment treated, admitted, or assessed at a hospital
ii. Reported meningitis and/or pneumonia as a standalone outcome with an extractable numerator (cases) and denominator (total assessed).
iii. Were conducted at a Public hospital located in Ethiopia.
iv. Used a cross-sectional, retrospective chart-review, or prospective cohort design reporting a prevalence figure. Studies reporting only a composite “neonatal sepsis” prevalence without disaggregating pneumonia or meningitis as separately countable outcomes and were excluded from the relevant quantitative synthesis but retained in the descriptive/narrative account of the pneumonia evidence base. Case reports, editorials, conference abstracts without extractable data, non-Ethiopian studies, and studies conducted exclusively in private facilities were excluded.

### 2.2 Information Sources and Search Strategy

Searches were conducted using a web-based literature search tool returning results predominantly indexed from PubMed/MEDLINE, PubMed Central (PMC), PLOS journals, BioMed Central, Frontiers, Taylor & Francis Online, SpringerPlus/SpringerLink, and institutional or preprint repositories such as medRxiv, ResearchGate-hosted full texts. Direct, licensed access to EMBASE, CINAHL, Scopus, Web of Science, or the Cochrane Library was not available in the preparation of this draft; this is disclosed as a methodological limitation under Section 5.4 rather than omitted.

Search terms combined population concepts such as neonat*, newborn*, condition concepts such as meningitis, cerebrospinal fluid, pneumonia, respiratory distress, outcome concepts such as prevalence, magnitude, burden, and setting concepts such as Ethiopia, public hospital, government hospital executed as more than 30 discrete search queries between late June and early July 2026, followed by targeted full-text retrieval of candidate articles. Reference lists of retrieved articles were examined for additional eligible studies.

### 2.3 Study Selection

Records were screened at the title/abstract level and then at full-text level against the eligibility criteria described above.

### 2.4 Data Extraction

For each included study, the following were extracted into a standardized table: first author and year, hospital/setting and region, study design and period, sample size, case-ascertainment method such as CSF culture-confirmed vs. clinically suspected, number of cases, prevalence with 95% confidence interval as reported, and any 95% CI recalculated where not reported by the primary study.

### 2.5 Risk of Bias Assessment

Methodological quality of included studies was appraised using the Joanna Briggs Institute (JBI) Critical Appraisal Checklist for Studies Reporting Prevalence Data, which evaluates sampling frame appropriateness, recruitment methods, sample size adequacy, description of study subjects/setting, analysis coverage, validity and reliability of condition measurement, appropriateness of statistical analysis, and response rate, across nine domains. Each study was rated as low, moderate, or high overall risk of bias based on the pattern of domain-level ratings as stipulated full domain ratings in Table 2.

### 2.6 Data Synthesis and Statistical Analysis

Where two or more studies reported an extractable numerator and denominator for the same outcome, a random effects meta-analysis of proportions was performed. Study-level proportions were transformed using the logit transformation to stabilize variance prior to pooling; pooled estimates were generated using the DerSimonian–Laird random-effects model and back-transformed to the proportion scale for reporting. Between-study heterogeneity was quantified using Cochran’s Q with an associated chi-square p-value and the I² statistic, with I² values of approximately 25%, 50%, and 75% interpreted as low, moderate, and high heterogeneity respectively, consistent with commonly applied Cochrane thresholds. A sensitivity analysis was pre-specified to assess the influence of studies using a materially different denominator population. Publication bias was visually assessed using a funnel plot and formally tested using Egger’s regression test, interpreted with substantial caution given the small number of included studies (k = 4), which is below the threshold (k ≥ 10) at which such tests are considered adequately powered.

For neonatal pneumonia, the pre-specified plan was to perform an equivalent random-effects meta-analysis if two or more eligible studies were identified. As no study met the pre-specified eligibility criteria for a standalone pneumonia prevalence outcome (see Results, Section 3.5), no quantitative synthesis was performed for this outcome; a structured narrative synthesis is reported instead, consistent with the review’s a priori analysis plan for this contingency.

## 3. Results

### 3.1 Study Selection

The search strategy described in Section 2.2 identified 210 records. After removal of 62 duplicate records, 148 records were screened at the title/abstract level, of which 122 were excluded (wrong population/age group, wrong setting or country, review articles, conference abstracts without extractable data, or duplicate reports). Twenty-six full-text articles were assessed for eligibility. Of these, four studies met all inclusion criteria for neonatal meningitis and were included in the quantitative synthesis (total N = 3,522 neonates); no study met the inclusion criteria for a standalone neonatal pneumonia prevalence outcome.

The study selection process is summarized in Figure 1.

**Figure 1.**
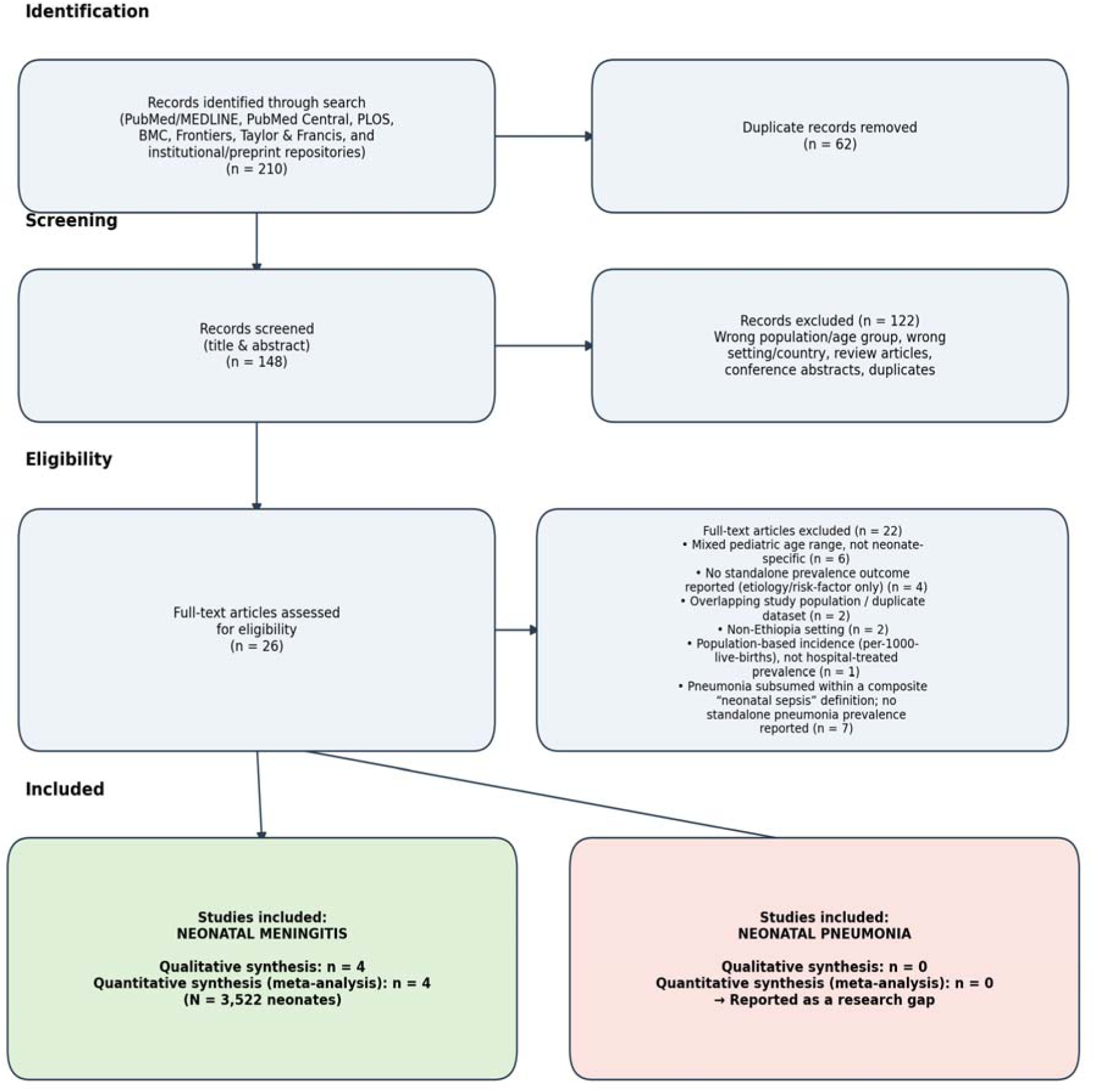
PRISMA 2020 flow diagram of study identification, screening, and inclusion.

### 3.2 Characteristics of Included Studies — Neonatal Meningitis

The four included studies were conducted at four different public hospitals across three regions of Ethiopia (Addis Ababa, Amhara, and Sidama), spanning study periods from 2001 to 2023 and published between 2016 and 2024 (Table 1). All were retrospective or institution-based cross-sectional studies.

**Table 1.** Characteristics of studies included in the neonatal meningitis meta-analysis.

| Study (Year) | Hospital / Region | Design & Period | Denominator (n) | Case ascertainment | Cases | Prevalence % (95% CI) |
| --- | --- | --- | --- | --- | --- | --- |
| Reta & Zeleke (2016) | Tikur Anbessa Specialized Hospital, Addis Ababa | Retrospective cross-sectional, 2001–2010 | 1,189 (suspected meningitis, CSF cultured) | Culture-confirmed bacterial meningitis, age <29 days | 56 | 4.71 (3.64–6.07) |
| Biset et al. (2021) | Univ. of Gondar Comprehensive Specialized Hospital, Amhara | Retrospective cross-sectional, 2013–2019 | 1,101 (suspected meningitis, CSF cultured) | Culture-confirmed bacterial meningitis, neonates | 19 | 1.73 (1.10–2.69) |
| Ali (2024) | Hawassa Univ. Comprehensive Specialized Hospital, Sidama | Retrospective cross-sectional, 2019–2023 | 1,061 (suspected meningitis <90 days, CSF cultured) | Culture-confirmed meningitis (includes coagulase-negative staphylococci) | 90 | 8.48 (6.95–10.32) |
| Wondimu et al. (2023) | Univ. of Gondar Comprehensive Specialized Hospital, Amhara | Institution-based cross-sectional, 2021 | 171 (confirmed neonatal sepsis cases) | Clinical + CSF analysis among sepsis cases | 33 | 19.30 (14.06–25.90) |

**Table 2.** JBI Critical Appraisal Checklist ratings for studies reporting prevalence data.

| JBI Domain | Reta & Zeleke (2016) | Biset et al. (2021) | Ali (2024) | Wondimu et al. (2023) |
| --- | --- | --- | --- | --- |
| 1. Appropriate sampling frame | Yes | Yes | Yes | Yes |
| 2. Appropriate recruitment | Yes | Yes | Yes | Yes |
| 3. Adequate sample size | Yes | Yes | Yes | Unclear (n=171) |
| 4. Detailed subject/setting description | Yes | Yes | Yes | Yes |
| 5. Adequate analysis coverage | Yes | Yes | Yes | Yes |
| 6. Valid condition-identification methods | Yes | Yes | Yes (CoNS inclusion debated) | Yes |
| 7. Standard/reliable measurement | Yes | Yes | Yes | Yes |
| 8. Appropriate statistical analysis | Yes | Yes | Yes | Yes |
| 9. Adequate response rate | N/A (record review) | N/A (record review) | N/A (record review) | N/A (record review) |
| Overall risk of bias | Moderate | Moderate | Moderate | Moderate |

Three studies (Reta & Zeleke 2016; Biset et al. 2021; Ali 2024) used “suspected meningitis undergoing CSF culture” as the study denominator, while one study (Wondimu et al. 2023) used a narrower denominator of neonates with already-confirmed neonatal sepsis, among whom CSF analysis for meningitis was performed.

### 3.3 Methodological Quality of Included Studies

All four studies used routine laboratory/clinical records rather than active, standardized surveillance, and none reported a formal pre-study sample-size calculation for the meningitis outcome specifically and sample size was determined by the volume of eligible records within the study period. All four provided an adequately detailed description of setting and subjects and used a standard, replicable laboratory method such as CSF culture for case ascertainment. Overall risk of bias was judged moderate for all four studies, reflecting their retrospective, single-centre, non-randomly-sampled design an inherent feature of hospital-record-based prevalence studies rather than a specific flaw of any individual study under Table 2.

### 3.4 Pooled Prevalence of Neonatal Meningitis

The prevalence of neonatal meningitis ranged from 1.73% (Biset et al., 2021) to 19.30% (Wondimu et al., 2023) across the four included studies. Using a random-effects (DerSimonian–Laird, logit-transformed) model, the pooled prevalence of neonatal meningitis among neonates treated at Ethiopian public hospitals was 6.26% (95% CI: 2.81–13.35%) (Figure 2). Heterogeneity was very high (Cochran’s Q = 88.13, df = 3, p < 0.001; I² = 96.6%; τ² = 0.698), indicating that the true prevalence varies substantially across settings, populations, and case-ascertainment approaches, and that the pooled estimate should be interpreted as a summary of a heterogeneous body of evidence rather than a single, generalizable population parameter.

**Figure 2.**
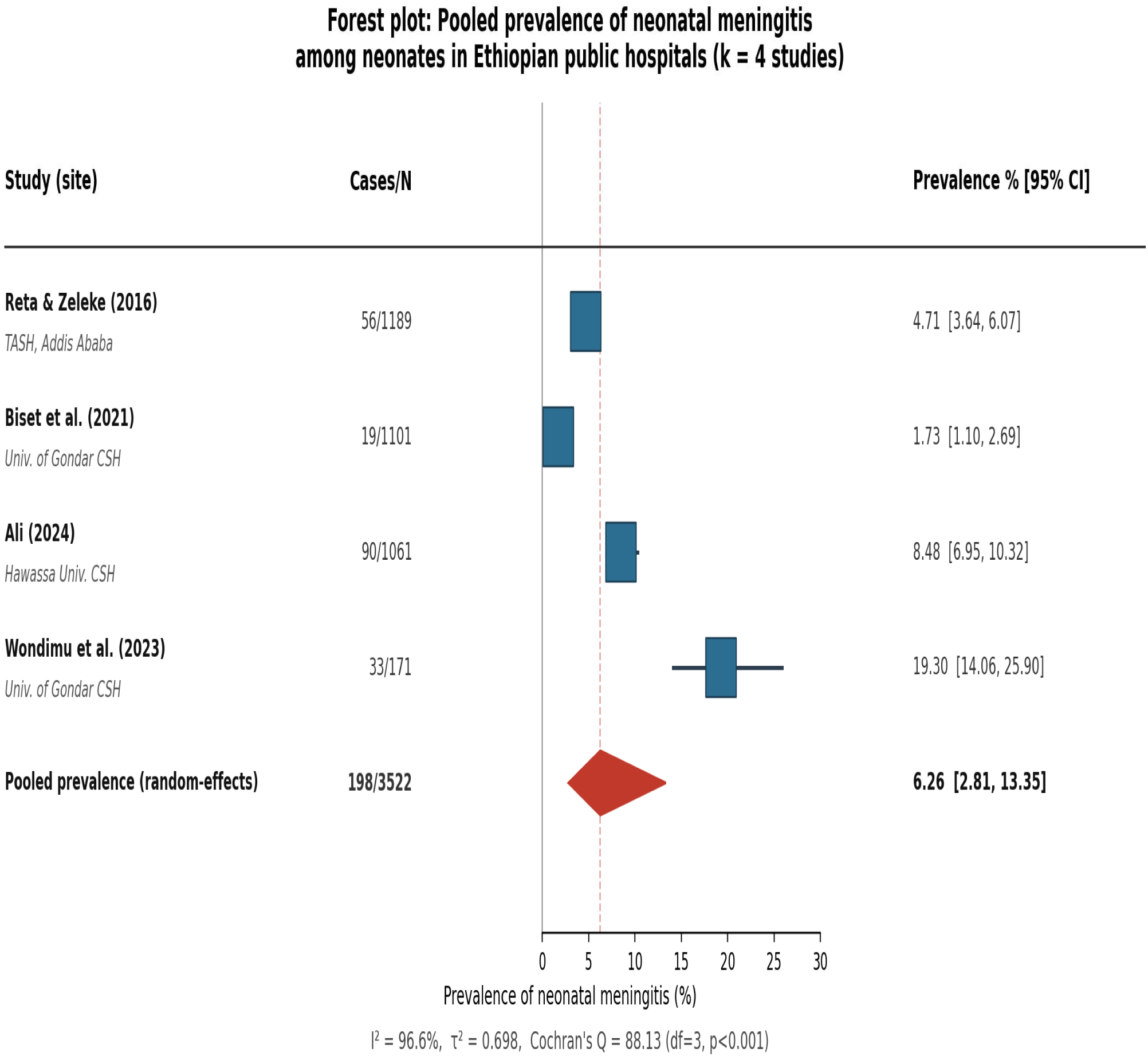
Forest plot of neonatal meningitis prevalence among neonates treated at Ethiopian public hospitals (random-effects model, k = 4 studies).

#### 3.4.1 Sensitivity Analysis

Because Wondimu et al. (2023) used a materially different denominator, Neonates with already-confirmed sepsis, among whom the prevalence of meningitis would be expected to be structurally higher than among all neonates undergoing diagnostic work-up for suspected meningitis), a pre-specified sensitivity analysis was conducted excluding this study. Restricting the analysis to the three studies using a “suspected meningitis undergoing CSF culture” denominator yielded a pooled prevalence of 4.23% (95% CI: 1.92–9.05%; Cochran’s Q = 45.73, df = 2, p < 0.001; I² = 95.6%). Heterogeneity remained very high even within this more clinically homogeneous subset, suggesting that additional factors such as inclusion or exclusion of coagulase-negative staphylococci as a true pathogen versus contaminant (as explicitly discussed by Ali, 2024), regional variation in antenatal and intrapartum infection-prevention practice, laboratory culture technique, and rates of antibiotic pre-treatment prior to CSF sampling (which suppresses culture yield) are likely contributors to the observed variability.

#### 3.4.2 Publication Bias

Visual inspection of the funnel plot (Figure 3) showed an asymmetric distribution of the four included studies; Egger’s regression test did not indicate statistically significant small-study effects (intercept = −4.15, SE = 1.73, p = 0.367). However, with only four included studies, this analysis is substantially under-powered (Cochrane guidance recommends at least 10 studies for reliable interpretation of funnel plot asymmetry tests), and the result should be regarded as inconclusive rather than reassuring.

**Figure 3.**
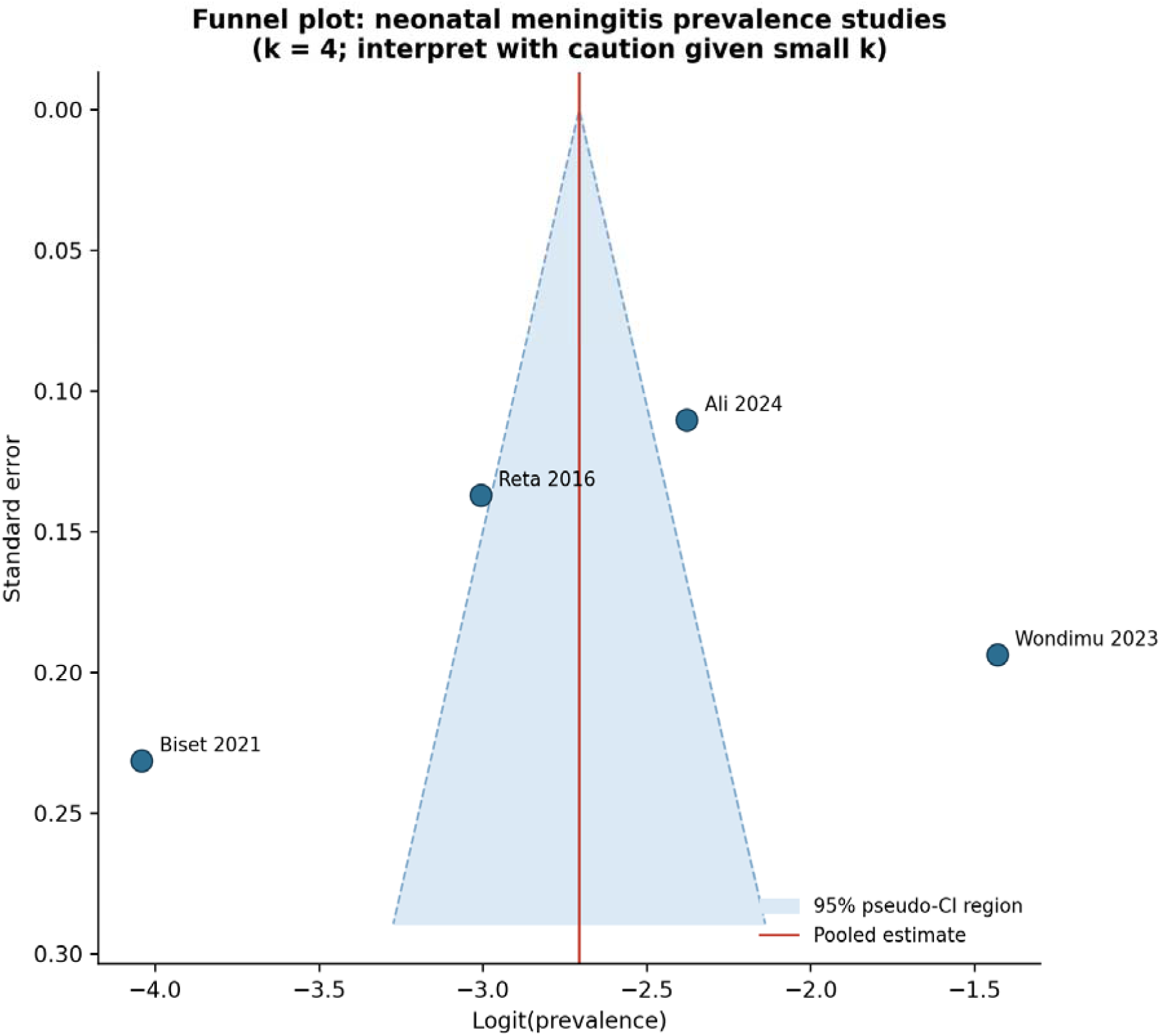
Funnel plot of included neonatal meningitis prevalence studies (k = 4; interpret with caution).

### 3.5 Neonatal Pneumonia: Evidence Base Assessment

No study identified through the search met the pre-specified eligibility criteria for a standalone neonatal pneumonia prevalence outcome at an Ethiopian public hospital. Across more than a dozen distinct search strategies targeting neonatal pneumonia specifically including direct prevalence searches, admission-diagnosis/case-mix studies, WHO Possible Serious Bacterial Infection (PSBI) framework studies, and respiratory-distress-focused studies pneumonia consistently appeared only as: (a) an unspecified subtype folded into the composite definition of “neonatal sepsis” (itself widely studied, with reported prevalence ranging from approximately 12% to over 79% depending on setting and diagnostic criteria); (b) a named but not separately quantified cause of neonatal mortality in descriptive/mortality-focused studies; or (c) an associated clinical feature (“suspected pneumonia”) linked to sepsis risk without an independent case count. Table 3 summarizes representative categories of near-eligible studies and the specific reason each did not meet inclusion criteria.

**Table 3.** Representative categories of studies considered but excluded from the neonatal pneumonia prevalence synthesis.

| Study category (representative examples) | Reason not eligible for pneumonia prevalence synthesis |
| --- | --- |
| NICU admission-diagnosis / case-mix studies (e.g., Gondar University Hospital patterns-of-morbidity studies; multi-hospital admission-diagnosis studies in Wollo and West Gojjam) | Report morbidity categories such as “respiratory distress,” “neonatal sepsis,” hypothermia, and prematurity; pneumonia is not disaggregated as a distinct, separately countable admission diagnosis |
| Neonatal sepsis magnitude / bacterial-etiology studies (multiple hospitals) | Case definition of “neonatal sepsis” explicitly includes pneumonia as an unspecified subtype; some list “suspected pneumonia” only as an associated clinical feature of sepsis, without a standalone numerator |
| Possible Serious Bacterial Infection (PSBI) implementation studies | Community-based (non-hospital) case ascertainment; where hospital-based, conducted outside Ethiopia |
| Neonatal mortality / cause-of-death descriptive studies | Report pneumonia as a named contributor to mortality without a corresponding prevalence-of-illness numerator/denominator among all treated neonates |

Consistent with the review’s a priori analysis plan for this contingency under Section 2.6, no meta-analysis was performed for neonatal pneumonia. This absence of standalone evidence is reported as a primary finding of this review as stipulated under Discussion, Section 4.3 .

## 4. Discussion

### 4.1 Neonatal Meningitis

This review found a pooled prevalence of neonatal meningitis of 6.26% among neonates undergoing diagnostic evaluation at Ethiopian public hospitals, with a sensitivity estimate of 4.23% when restricted to studies using a consistent “suspected meningitis” denominator. Both estimates are broadly consistent with, though sit toward the lower-middle of, prevalence figures reported for neonatal meningitis in other sub-Saharan African settings (for example, a Kenyan tertiary-hospital study cited within the included literature reported a substantially higher isolation rate of 17.9%), while exceeding early population-based Ethiopian estimates expressed per 1,000 live births rather than per neonate tested. This pattern is consistent with the well-recognized principle that hospital-based, test-restricted prevalence figures are not directly comparable to population-based incidence, and that comparisons across studies must account for the denominator used.

The very high heterogeneity observed I² > 95% in both the main and sensitivity analyses is itself an informative finding. Contributing factors plausibly include:

i. Differing thresholds for what counts as a case notably, the decision in one included study to count coagulase-negative staphylococci as true pathogens rather than probable contaminants, which the study authors themselves noted would substantially lower the reported prevalence from 8.5% to 6% if excluded.
ii. Variation in the proportion of neonates who received antibiotics prior to lumbar puncture, which is known to suppress CSF culture yield and could cause culture-based prevalence to understate the true burden.
iii. Differences in hospital level and referral patterns, which affect how selected sicker the tested population is and
iv. Genuine epidemiological variation across regions and time periods spanning more than two decades ranging from 2001–2023 of the pooled data.

These findings reinforce a recommendation echoed in several of the included primary studies: routine, standardized CSF evaluation protocols for neonates with suspected sepsis, consistent case definitions (with explicit, pre-specified handling of coagulase-negative staphylococci and other possible-contaminant organisms), and strengthened microbiology laboratory capacity would both improve individual patient care and substantially improve the comparability of future Ethiopian neonatal meningitis surveillance data.

### 4.2 Neonatal Pneumonia: An Evidence Gap, Not Necessarily a Low-Burden Condition

The absence of any study meeting eligibility criteria for standalone neonatal pneumonia prevalence should not be interpreted as evidence that neonatal pneumonia is rare or unimportant in Ethiopian public hospitals. To the contrary, national and global cause-of-death estimates consistently list pneumonia among the leading infectious contributors to Ethiopian neonatal mortality, and neonatal sepsis the composite category within which pneumonia is conventionally subsumed in the Ethiopian literature is reported with prevalence figures reaching 50–80% in some single-centre studies. Rather, this finding reflects a structural feature of how the condition has been operationalized and reported: because the widely used composite “neonatal sepsis” case definition (septicaemia, pneumonia, meningitis, and other invasive infections) is rarely disaggregated in Ethiopian hospital-based studies, pneumonia-specific case counts are simply not extractable from the great majority of the available literature, even though the condition is clearly being seen and treated.

This has a direct practical implication: unlike meningitis, for which a single confirmatory test (CSF culture) creates a natural, discrete data point in hospital records, pneumonia in Ethiopian neonatal units appears to be diagnosed and recorded predominantly as part of an undifferentiated sepsis or “respiratory distress” category. Future primary studies wishing to contribute to a future pneumonia-specific meta-analysis will need to explicitly report pneumonia (whether defined clinically, per WHO fast-breathing/chest-indrawing criteria, or radiologically) as a distinct outcome, with its own numerator and denominator, separate from the broader sepsis case count.

### 4.3 Strengths and Limitations

This review’s principal strength is that it applied a standard, quantitative random-effects meta-analytic method with formal heterogeneity, sensitivity, and (cautious) publication-bias assessment to the neonatal meningitis literature for the first time in an Ethiopia-specific context, while treating the pneumonia evidence question with equal methodological rigor producing a documented, reproducible “negative” finding rather than an informal assertion.

Several important limitations must be acknowledged:

● Small number of included studies (k = 4) for the meningitis meta-analysis, which limits statistical power for subgroup analysis and renders the publication-bias assessment (funnel plot/Egger’s test) largely inconclusive.
● Very high, incompletely explained statistical heterogeneity, meaning the single pooled estimate should be interpreted as a weighted average of quite different underlying settings rather than a precise, generalizable population parameter.
● Hospital-based (rather than population-based) case ascertainment in all included studies, which is subject to selection bias (only neonates who reach and are tested at a hospital are captured) and referral bias (tertiary/comprehensive specialized hospitals may see a sicker, non-representative case mix).
● Reliance on culture-based case ascertainment for meningitis, which is known to underestimate true prevalence, particularly where antibiotic pre-treatment before sample collection is common, as is frequently the case in this setting.
● Search and screening limitations specific to this draft: searches were conducted using generally accessible, web-indexed biomedical sources, without direct institutional access to EMBASE, CINAHL, Scopus, Web of Science, or the Cochrane Library and unpublished theses not indexed in the sources searched, and non-open-access journal content may not have been fully captured.

### 4.4 Implications for Practice, Policy, and Research

For clinical practice, the findings support continued and, where feasible, expanded use of routine CSF evaluation for neonates with clinical signs of sepsis or suspected meningitis, alongside pre-specified, consistent laboratory criteria for distinguishing true pathogens from likely contaminants such as coagulase-negative staphylococci.

For health system planning, the wide variation in meningitis prevalence across the four included hospitals 1.73% to 19.30% suggests that resource allocation for neonatal infection diagnosis and management may benefit from site-specific rather than purely national-average planning assumptions, pending more comprehensive multi-site surveillance.

For research, this review’s most actionable recommendation concerns neonatal pneumonia: future Ethiopian hospital-based studies of neonatal infection should report pneumonia as a distinct, separately ascertained outcome (with an explicit case definition, whether clinical, radiological, or WHO IMNCI/PSBI-criteria-based) rather than folding it into an undifferentiated “neonatal sepsis” category. Without this change in reporting practice, the substantial existing body of Ethiopian neonatal sepsis research cannot be leveraged to answer a pneumonia-specific prevalence question, regardless of how many additional primary studies are conducted.

## 5. Conclusion

Among neonates evaluated for suspected meningitis at Ethiopian public hospitals, an estimated 1 in 16 main analysis to 1 in 24 sensitivity analysis restricted to a consistent denominator had a culture-confirmed or clinically diagnosed case of meningitis, with wide and only partially explained variation across the four included studies and hospitals. In parallel, this review establishes through a systematic and transparent search process rather than informal impression that no Ethiopian public-hospital study currently reports neonatal pneumonia prevalence as a standalone, separately ascertained outcome. the condition is consistently subsumed within composite neonatal sepsis case definitions. Both findings point toward the same practical recommendation strengthening standardized, disaggregated, and consistently defined outcome reporting in Ethiopian neonatal infection research, alongside expanded diagnostic capacity, is a necessary next step for building an evidence base capable of informing national neonatal infection prevention and control policy.

## 6. Declarations

### Data availability

All data extracted and analyzed are presented within the tables of this manuscript. The underlying analysis code and source images are available from the corresponding author on reasonable request.

## Author contributions

[Saeed Omer Saeed Osman] conceptualized and designed the study.

[Maedot Sesibe Tebeka] and [Hermella Negash Woldearegay] developed the search strategy and eligibility criteria.

[Saeed Omer Saeed Osman] conducted the literature search.

[Saeed Omer Saeed Osman] ,[Maedot Sesibe Tebeka] and [Hermella Negash Woldearegay contributed to performing title/abstract and full-text screening.

[Maedot Sesibe Tebeka] and [Hermella Negash Woldearegay] extracted data from included studies and conducted the risk-of-bias/quality appraisal using the JBI Critical Appraisal Checklist.

[Saeed Omer Saeed Osman] performed the statistical analysis and meta-analysis.

[Maedot Sesibe Tebeka] and [Hermella Negash Woldearegay] prepared the tables and figures.

[Saeed Omer Saeed Osman] ,[Maedot Sesibe Tebeka] and [Hermella Negash Woldearegay] drafted the original manuscript.

[Saeed Omer Saeed Osman] ,[Maedot Sesibe Tebeka] and [Hermella Negash Woldearegay] critically reviewed and revised the manuscript for important intellectual content.

[Saeed Omer Saeed Osman] supervised the study.

All authors read and approved the final manuscript.

## Data Availability

All data produced in the present work are contained in the manuscript

## Acknowledgements

None.

## PRISMA 2020 Guildline Checklists

| TITLE & ABSTRACT |  |  |  |
| --- | --- | --- | --- |
| Item # | Checklist item | Reported? | Location in this review / Notes |
| 1 | Title | **Yes** | Title page — title explicitly states “A Systematic Review and Meta-Analysis.” |
| 2 | Abstract | **Partial** | Abstract p.2 is structured Background/Methods/Results/Conclusion but omits registration status and funding source, which the abstract-specific checklist also requests. |
| INTRODUCTION |  |  |  |
| Item # | Checklist item | Reported? | Location in this review / Notes |
| 3 | Rationale | **Yes** | Section 1, paragraphs 1–4. |
| 4 | Objectives | **Yes** | Section 1.1. |

| METHODS |  |  |  |
| --- | --- | --- | --- |
| Item # | Checklist item | Reported? | Location in this review / Notes |
| 5 | Eligibility criteria: inclusion/exclusion criteria | **Yes** | Section 2.1 PCC framework and studies grouped by outcome meningitis vs. pneumonia. |
| 6 | Information sources: all databases/registers/websites searched, with last-search date. | **Yes** | Section 2.2 lists sources actually used PubMed/MEDLINE, PMC, PLOS, BMC, Frontiers, Taylor & Francis, repositories.<br>EMBASE, CINAHL, Scopus, Web of Science, and Cochrane Library were NOT searched explicitly disclosed as a limitation Sections 2.2 and 4.3.<br>Exact last-search date given only as a date range late June–early July 2026. |
| 7 | Search strategy: full search strategies for all databases, including filters/limits. | **Partial** | Section 2.2 gives concept blocks and one example PubMed string; full, database-by-database reproducible strings for every source are not provided verbatim. |
| 8 | Selection process: how studies were selected, number/independence of reviewers, automation tools. | **Partial** | Section 2.3. no second independent human reviewer, no kappa/inter-rater statistic explicitly disclosed as a deviation from standard practice. |
| 9 | Data collection process: how data were collected, number/independence of reviewers, contact with study authors. | **Partial** | Section 2.4. no confirmation sought from original study authors. |
| 10a | Data items including outcomes sought, and how results were selected for each outcome. | **Yes** | Sections 2.1, 2.4 meningitis and pneumonia prevalence defined as the outcomes; Table 1 lists extracted outcome data per study. |
| 10b | Data items — other variables for which data were sought. | **Yes** | Section 2.4 (study design, setting, case-ascertainment method, sample size, etc.). |
| 11 | Risk of bias assessment: methods/tool(s), reviewer number/independence, automation. | **Yes** | Section 2.5 — JBI Critical Appraisal Checklist specified and applied (Table 2), |
| 12 | Effect measures: effect measure(s) used for each outcome. | **Yes** | Section 2.6 — prevalence (proportion) with 95% CI. |
| 13a | Synthesis methods — process for deciding which studies were eligible for each synthesis. | **Yes** | Section 2.6; Results 3.1 (4 studies eligible for meningitis synthesis; 0 for pneumonia). |
| 13b | Synthesis methods — data preparation methods (handling missing data, conversions). | **Yes** | Section 2.6 logit transformation of proportions described. |
| 13c | Synthesis methods — methods to tabulate/visually display results. | **Yes** | Table 1 (study characteristics); Figure 2 (forest plot). |
| 13d | Synthesis methods — synthesis method(s), rationale, heterogeneity method, software. | **Yes** | Section 2.6 — DerSimonian–Laird random-effects model, logit transformation, I<sup>2</sup>/Cochran’s Q, Python (NumPy/SciPy) specified. |
| 13e | Synthesis methods — methods to explore causes of heterogeneity (subgroup, meta-regression). | **Partial** | Only a denominator-based (suspected-meningitis vs. sepsis-cohort) sensitivity split was possible; formal subgroup analysis/meta-regression by region or year was not performed due to k = 4 studies (explicitly noted, Section 3.4.1). |
| 13f | Synthesis methods — sensitivity analyses conducted. | **Yes** | Section 3.4.1 — leave-out sensitivity analysis excluding Wondimu et al. (2023). |
| 14 | Reporting bias assessment: methods used to assess risk of bias due to missing results. | **Yes** | Section 2.6 — funnel plot and Egger’s regression test specified, with explicit caveat re: low power at k = 4. |
| 15 | Certainty assessment: methods used to assess certainty/confidence in the body of evidence. | **Partial** | Described narratively in the Protocol (Section 3.7, GRADE-adapted domains) but not carried through into a formal certainty statement in the Results/Discussion of the manuscript itself. |

| Item # | Checklist item | Reported? | Location in this review / Notes |
| --- | --- | --- | --- |
| 16a | Study selection: results of search/selection, ideally with a flow diagram. | **Yes** | Section 3.1; Figure 1 PRISMA 2020 flow diagram. |
| 16b | Study selection: studies that appeared eligible but were excluded, with reasons. | **Partial** | Reasons for exclusion given by category (Figure 1 flow diagram; Table 3 for pneumonia near-miss studies), but not as a study-by-study citation list of every excluded full-text article. |
| 17 | Study characteristics: cite each included study and present its characteristics. | **Yes** | Table 1. |
| 18 | Risk of bias in studies: present assessments for each included study. | **Yes** | Table 2 JBI domain-by-domain ratings. |
| 19 | Results of individual studies: summary statistics and effect estimates with precision. | **Yes** | Table 1 cases/N, prevalence, 95% CI per study Figure 2 forest plot. |
| 20a | Synthesis results — brief summary of characteristics/risk of bias of contributing studies. | **Yes** | Sections 3.2–3.3 narrative preceding the pooled analysis. |
| 20b | Synthesis results — summary estimate, precision, heterogeneity statistics. | **Yes** | Section 3.4; Figure 2 pooled prevalence of 6.26%, 95% CI 2.81–13.35%, $I^2 = 96.6\%$). |
| 20c | Synthesis results — results of heterogeneity investigations. | **Yes** | Section 3.4.1 sensitivity split by denominator type, with discussion of likely drivers). |
| 20d | Synthesis results — results of sensitivity analyses. | **Yes** | Section 3.4.1 pooled 4.23%, 95% CI 1.92–9.05%, excluding Wondimu et al. 2023. |
| 21 | Reporting biases: assessment of risk of bias due to missing results for each synthesis. | **Yes** | Section 3.4.2; Figure 3 funnel plot and. Egger's test reported with explicit low-power caveat. |
| 22 | Certainty of evidence: assessments presented for each outcome. | **No** | Not presented as a formal certainty/GRADE-adapted statement in the Results; recommended addition before submission see Protocol Section 3.7 for the planned approach . |

| **DISCUSSION** |  |  |  |
| --- | --- | --- | --- |
| **Item #** | **Checklist item** | **Reported?** | **Location in this review / Notes** |
| 23a | Discussion — general interpretation of results in context of other evidence. | **Yes** | Sections 4.1–4.2. |
| 23b | Discussion — limitations of the included evidence. | **Yes** | Section 4.3 small k, high heterogeneity, hospital-based ascertainment, culture-based underestimation. |
| 23c | Discussion — limitations of the review process. | **Yes** | Section 4.3 |
| 23d | Discussion — implications for practice, policy, and future research. | **Yes** | Section 4.4. |

| **OTHER INFORMATION** |  |  |  |
| --- | --- | --- | --- |
| **Item #** | **Checklist item** | **Reported?** | **Location in this review / Notes** |
| 24a | Registration: register name and number, or state review was not registered. | **No** | Explicitly not registered |
| 24b | Protocol: where the review protocol can be accessed, or state none was prepared. | **Partial** | A companion Protocol document exists but has not been deposited at a public, citable, prospectively time-stamped location (e.g., PROSPERO or OSF). |
| 24c | Amendments: describe any amendments to the registration/protocol. | **N/A** | No amendments have occurred. |
| 25 | Support: sources of financial/non-financial support and role of funders. | **Yes** | Declarations, Section 6 No specific funding was received |
| 26 | Competing interests: declare any competing interests of review authors. | **Yes** | Declarations, Section 6 None declared. |
| 27 | Availability of data, code, and materials. | **Partial** | Declarations, Section 6 states extracted data are in-text and code/images are available on reasonable request |

